# A Randomised, Double-Blind, Sham-Controlled Trial of Deep Brain Stimulation of the Bed Nucleus of the Stria Terminalis for Treatment-Resistant Obsessive-Compulsive Disorder

**DOI:** 10.1101/2020.10.24.20218024

**Authors:** Philip E. Mosley, François Windels, John Morris, Terry Coyne, Rodney Marsh, Andrea Giorni, Adith Mohan, Perminder Sachdev, Emily O’Leary, Mark Boschen, Pankaj Sah, Peter A. Silburn

## Abstract

Deep brain stimulation (DBS) is a promising treatment for severe, treatment-resistant obsessive-compulsive disorder (OCD). Here, nine participants (four females, mean age 47.9 ±10.7 years) were implanted with DBS electrodes bilaterally in the bed nucleus of the stria terminalis (BNST). Following a one-month postoperative recovery phase, participants entered a three-month randomised, double-blind, sham-controlled phase before a twelve-month period of open-label stimulation incorporating a course of cognitive behavioural therapy (CBT). The primary outcome measure was OCD symptoms as rated with the Yale-Brown Obsessive-Compulsive Scale (YBOCS). In the blinded phase, there was a significant benefit of active stimulation over sham (*p* = 0.025, mean difference 4.9 points). After the open phase, the mean reduction in YBOCS was 16.6 ±1.9 points (*Χ*^2^ (11) = 39.8, p = 3.8 × 10^−5^), with seven participants classified as responders. CBT resulted in an additive YBOCS reduction of 4.8 ±3.9 points (*p* = 0.011). There were two serious adverse events related to the DBS device, the most severe of which was an infection during the open phase necessitating device explantation. There were no psychiatric adverse events related to stimulation. An analysis of the structural connectivity of each participant’s individualised stimulation field isolated right-hemispheric fibres associated with YBOCS reduction. These included subcortical tracts incorporating the amygdala, hippocampus and stria terminalis, in addition to cortical regions in the ventrolateral and ventromedial prefrontal cortex, parahippocampal, parietal and extrastriate visual cortex. In conclusion, this study provides further evidence supporting the efficacy and tolerability of DBS for individuals with otherwise treatment-refractory OCD and identifies a connectivity fingerprint associated with clinical benefit.

## 2 INTRODUCTION

Obsessive-compulsive disorder (OCD) is a psychiatric condition with an estimated lifetime prevalence of between 1-2 % (Kessler *et al*., 2005). It is characterised by the intrusion of ego-dystonic, anxiety-provoking thoughts (obsessions). These are accompanied by mental acts or behaviours (compulsions), which must be carried out to neutralise the obsessions, or to mitigate anxiety associated with them (American Psychiatric Association., 2013). Remission of symptoms with pharmacological treatment is rare (Erzegovesi *et al*., 2001) and persistent impairment is relatively common even with combination therapy (Bloch *et al*., 2006). Psychological treatment is often intolerable for those with a severe illness: deliberate exposure to obsessive thoughts during cognitive behavioural therapy (CBT) is aversive and distressing (Issakidis and Andrews, 2002). These factors mean that OCD is a chronic disorder with a detrimental effect on functioning across the lifespan, making it a leading neuropsychiatric cause of global disability (Mathers *et al*., 2008).

Deep brain stimulation (DBS) is a reversible and adjustable form of targeted neuromodulation that has been used successfully for the treatment of movement disorders such as Parkinson’s disease for over 25 years (Benabid *et al*., 1994; Schuepbach *et al*., 2013). DBS was first employed for the treatment of intractable OCD in the late 1990s (Nuttin *et al*., 1999), with initial surgical targeting in the anterior limb of the internal capsule (ALIC) informed by prior work using ablative neurosurgery (Nuttin *et al*., 2003). Further work reproduced these encouraging preliminary outcomes (Abelson *et al*., 2005; Farrand *et al*., 2018; Goodman *et al*., 2010; Greenberg *et al*., 2006), finding improved response with posterior migration of the target to the region of the caudal nucleus accumbens (NAcc) (Greenberg *et al*., 2010). The anteromedial segment of the subthalamic nucleus (STN) has also been a promising target for neuromodulation (Mallet *et al*., 2008). More recently, two randomised, placebo-controlled, crossover trials of DBS at the NAcc/ALIC interface (Denys *et al*., 2010) and the BNST/ALIC interface (Luyten *et al*., 2016) demonstrated a statistically-significant benefit of active stimulation over sham.

The clinical benefits (and side effects) of DBS for movement disorders arise not only from the effect of focal stimulation at the target nucleus, but also from the modulation of distributed brain networks structurally and functionally connected to the stimulation field (Accolla *et al*., 2016; Akram *et al*., 2017; Chen *et al*., 2018; Horn *et al*., 2017; Mosley *et al*., 2020a; Vanegas-Arroyave *et al*., 2016). In a similar manner, brain networks associated with response to DBS for OCD can be delineated. In prior work, reduction in OCD symptoms 12-months after NAcc/ALIC DBS was associated with connectivity of the stimulation site with the right ventrolateral prefrontal cortex, with a fibre tract predictive of symptom reduction identified in the ventral ALIC bordering the BNST (Baldermann *et al*., 2019). A randomised trial directly comparing ALIC and anteromedial STN stimulation found both to be clinically effective targets but with distinct structural connectivity profiles and dissociable effects on mood and cognitive flexibility (Tyagi *et al*., 2019). However, a pooled analysis of four cohorts employing either STN or ALIC stimulation identified a universal tract associated with clinical response that could predict outcome in an out-of-sample cross-validation (Li *et al*., 2019). This tract traversed both the anteromedial STN and ventral ALIC, projecting to ventrolateral prefrontal cortex. Overall, these findings suggest that different surgical targets may act to reduce OCD symptoms through modulation of a shared network, whilst change amongst more fine-grained behavioural endophenotypes may result from modulation of networks that are not shared between targets (Dougherty, 2019).

In this study, using a randomised, double-blind, sham-controlled, staggered-onset design, we investigate the effects of DBS at the BNST/NAcc interface in a sample of Australian participants with severe, treatment-resistant OCD. We delineate the structural connectivity profile of effective stimulation and compare this with the aforementioned prior work. We also add CBT incorporating exposure and response prevention (ERP) to the open phase of the trial, in order to investigate whether this is now tolerable for our participants and leads to an additive clinical response, as has been identified in a previous cohort (Mantione *et al*., 2014). We report outcomes during the blinded phase and after one year of open stimulation following completion of CBT.

## 3 MATERIALS AND METHODS

### 3.1 Participants

All procedures were carried out in accordance with the experimental protocol approved by the Human Research Ethics Committees of the University of Queensland and UnitingCare Health. Participants aged 18-70 with severe, treatment-resistant OCD of at least five years duration were referred by their treating psychiatrists and evaluated independently by two psychiatrists in the research team (PEM and RM). The diagnosis of OCD was confirmed according to criteria defined by the Diagnostic and Statistical Manual of Mental Disorders, fifth edition (DSM-V) (American Psychiatric Association., 2013). Severity was denoted by a mean score of at least 24 on the Yale-Brown Obsessive-Compulsive Scale (YBOCS) (Goodman *et al*., 1989), measured twice at least two weeks apart by separate investigators. Treatment refractoriness was defined by insufficient response to at least: i) two trials of selective serotonin reuptake inhibitors at maximum tolerated dose for at least 12 weeks, ii) one trial of clomipramine at maximum tolerated dosage for at least 12 weeks, plus iii) one augmentation trial with an antipsychotic for at least eight weeks in combination with one of the aforementioned drugs, plus iv) one complete trial of ERP-based CBT confirmed by a psychotherapist. Exclusion criteria included pregnancy, a past history of a chronic psychotic or bipolar disorder, severe personality disorder, suicidality in the previous 12 months, substance use disorder (except tobacco), major neurological comorbidity or severe head injury, prior ablative neurosurgery and current implanted cardiac pacemaker, defibrillator or other neurostimulator. Suitable and consenting candidates were approved by an independent Mental Health Review Tribunal prior to neurosurgery. Prior to implantation of the first participant, the trial was registered on the Australian and New Zealand Clinical Trials Registry (Universal Trial Number: U1111-1146-0992).

### 3.2 Device Implantation

Bilateral implantation of Medtronic (Minneapolis, USA) 3389 quadripolar electrodes took place in a single-stage procedure using a Leksell stereotactic apparatus based on preoperative structural magnetic resonance neuroimaging (Supplementary Materials). The most ventral contact was sited posterior and inferior to the NAcc in the region of the lateral hypothalamus, with more dorsal contacts within the BNST approaching the posterior border of the NAcc.

Postoperative lead placement was confirmed with CT imaging. Electrodes were connected to an Activa PC+S implantable pulse generator (IPG) in either the pectoral or abdominal fascia. Analysis of long-term, ambulatory electrophysiological data will be reported in forthcoming work.

### 3.3 Timeline, Assessment and Intervention

Following device implantation, participants entered a one-month recovery phase during which all stimulators were off. Thereafter, participants began a three-month period during which their stimulators were either turned on or remained switched off whilst both participants and assessors were blinded to status. After this, participants continued in an open-label (unblinded) trial where all stimulators were on. Assessments took place at baseline one week before surgery, fortnightly in the recovery phase, monthly in the blinded phase and monthly for the first three months of the open phase, with the time between assessments subsequently extending to two and then three months. The primary outcome measure was OCD severity as assessed by the YBOCS score, derived from a ten-item semi-structured interview assessing obsessions and compulsions, with a maximum score of 40. Depressive symptoms were assessed as a secondary outcome with the Montgomery Åsberg Depression Rating Scale (MADRS) score, derived from a ten-item semi-structured interview with a maximum score of 60 (Montgomery and Asberg, 1979; Williams and Kobak, 2008). Participants were referred for a ten-session course of ERP-based CBT with a clinical psychologist (EOL or MB) during the open phase once DBS parameters had been optimised and YBOCS reduction had plateaued.

Stimulation was commenced in an identical manner for participants regardless of whether they were turned on in either the blinded or open-label phase. Contact 1 (left hemisphere) and contact 9 (right hemisphere) were selected with an initial stimulation amplitude of 1 Volt, a pulse-width of 90 microseconds and a frequency of 130 Hertz. Stimulation was increased at weekly to fortnightly intervals in increments of 0.5-1 Volt to a target of 4.5 Volts. Stimulation settings were symmetric between hemispheres. If there was a relative lack of response as assessed with the YBOCS, additional stimulation changes were trialled: including further increases in amplitude in 0.1 Volt increments, a trial of a pulse-width of 120 microseconds or the activation of a second contact on each electrode. Psychotropic medications were unchanged throughout the trial unless requested for clinical reasons by the participant’s usual psychiatrist.

### 3.4 Randomisation and Blinding

Participants were randomly allocated in a 1:1 ratio to ‘on’ or ‘off’ groups in the blinded phase by an external statistician, using an online tool (https://www.sealedenvelope.com). Only the lead neurologist (PAS) and programming psychiatrist (PEM) were informed of the allocation. The psychiatrist assessing primary and secondary outcomes (RM) remained blinded to participant status. To reduce the likelihood of participants becoming unblinded by sensations associated with active stimulation, no contact testing was performed and the slow titration protocol was followed in all cases.

### 3.5 Statistical Analysis

Data analysis was performed in the R software environment (R Core Team, 2014). In the blinded phase of the trial, the mean change in YBOCS and MADRS score was compared between groups with a two-sample *t*-test. After one year of open stimulation and following a course of CBT, the reduction in YBOCS and MADRS score was assessed with the package *lmerTest* (Kuznetsova *et al*., 2017) using a random-intercept, random-slope, linear mixed-effects model incorporating demographic variables and baseline severity

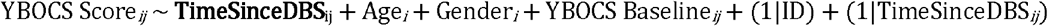

with _*i*_ denoting participant and _*j*_ denoting timepoint and the term in bold (the accrued effect of DBS over time on obsessive and depressive symptoms) being the coefficient of interest. Hypothesis testing on a null model (omitting TimeSinceDBS) was performed with the *anova* function in the *lavaan* package.

Consistent with prior work, participants were defined as responders for OCD and depression if they attained a reduction of 35 % in YBOCS score and 50 % in MADRS score respectively.

### 3.6 Electrode Localisation & Volume of Tissue Activation

DBS electrodes were localized using the Lead-DBS toolbox version 2.2 (https://github.com/netstim/leaddbs/tree/develop) (Horn and Kuhn, 2015; Horn *et al*., 2019). Preoperative structural acquisitions were co-registered with postoperative CT imaging and then normalized into common ICBM 2009b nonlinear asymmetric space using the SyN approach implemented in advanced normalization tools (ANTs) (Avants *et al*., 2008). Electrode trajectories were reconstructed after correcting for brainshift in postoperative acquisitions by applying a refined affine transform in a subcortical area of interest calculated pre- and postoperatively. For each electrode, a volume of activated tissue (VAT) was estimated using a volume conductor model of the DBS electrode and surrounding tissue, based on each participant’s individualised stimulation settings and a finite element method to derive the gradient of the potential distribution (Horn *et al*., 2019). An electric field (E-field) distribution was also modelled (Vorwerk *et al*., 2018).

### 3.7 Structural Connectivity Estimation and YBOCS Reduction

Three methods were used to assess the relationship between the structural connectivity of the stimulation field and the primary outcome measure. Firstly, using the Lead-DBS toolbox, each participant’s VAT in each hemisphere was integrated with a normative whole-brain structural connectome incorporating six million fibres derived from 985 participants in the Human Connectome Project who had undertaken multi-shell diffusion-weighted imaging (Van Essen *et al*., 2013). Fibres traversing each participant’s VAT were selected from the group connectome based on the E-field gradient strength (i.e. fibres in peripheral VAT regions with a low E-field were down-weighted) and projected to the volumetric surface of the ICBM 2009b nonlinear asymmetric brain in 1 mm isotropic resolution. A connectivity profile for each participant was expressed as the weighted number of fibre tracts between the stimulation site and each brain voxel. Subsequently, each voxel on the corresponding connectivity profile was correlated with clinical improvement on the YBOCS score using a Spearman rank correlation coefficient, forming an ‘R-map’. Combined across all participants, these maps identify regions to which strong connectivity is associated with good clinical outcome, modelling ‘optimal’ connectivity from the stimulation field to the rest of the brain (Horn *et al*., 2017). To verify these findings, the data were cross-validated in a leave-one-out design. Each participant was sequentially excluded and the optimal connectivity profile was computed on the remaining participants. Subsequently, YBOCS reduction was predicted for the excluded participant based on comparison between individual and group connectivity estimates (using a Fisher z-transformed spatial correlation coefficient) and the empirical outcome was correlated with the predicted outcome derived from the remaining sample.

Secondly, individual fibres associated with YBOCS reduction were identified. Each whole-brain fibre was tested across the cohort between participants with a stimulation volume that encompassed the fibre (connected) and those where the fibre did not traverse the volume (unconnected). If there was a significant difference between YBOCS reduction in participants with connected and unconnected VATs (using a two-sided, two-sample t-test), then this fibre was identified as discriminative of outcome. This process yielded a ‘fibre t-score’, with high-values indicating that this fibre was strongly discriminative of clinical outcome (Baldermann *et al*., 2019). Only the top 5 % of fibres positively correlated with the primary outcome variable were selected for analysis to mitigate the risk of false positive associations.

Finally, to explore whether connectivity to specific cortical regions was related to YBOCS reduction, a region of interest analysis was informed by findings from the aforementioned methods. Cortical parcellations were derived from the Desikan-Killiany-Tourville labelling protocol (Klein and Tourville, 2012; Klein *et al*., 2017), with connectivity estimates between each VAT and cortical region entered into the multivariate linear mixed-effects model to derive an estimate of effect size and statistical significance.

### 3.8 Data Availability

A de-identified data set containing demographic and outcome data can be provided by Dr Philip Mosley on application, subject to institutional review board approval. Local ethics caveats and clinical privacy issues prohibit sharing of individual imaging data but a copy of the Lead-DBS group analysis database can be supplied.

## 4 RESULTS

### 4.1 Participants

Nine participants (four females, mean age 47.9 ±10.7 years, mean baseline YBOCS 32.7 ±2.6) were recruited, randomised and implanted (Figure 1 and Table 1). The first participant was implanted in late 2015 and the last in early 2019. Contacts selected for activation were located in the BNST posterior to the NAcc and inferomedial to the ventral pallidum (Figure 2). All participants completed the blinded phase and data was analysed according to originally-assigned group. During the open phase, one participant developed an IPG infection necessitating DBS device explantation and exit from the trial. Scores at trial exit were carried forward for the two remaining data points. The eight remaining participants completed a course of ERP-based CBT. One participant (06) switched antidepressants and antipsychotics during the trial due to non-response to DBS and persistence of clinically-significant symptoms.

**Table 1.** Details of Participants

| **Participant**<br>**Age §**<br>**Gender** | **OCD Phenomenology & Age of Onset**<br>**Previous Therapies † &**<br>**Comorbidities**<br>**YBOCS at Baseline** | **Psychotropic Medication**<br>**Chronic Stimulation**<br>**Parameters ‡** | **Percentage YBOCS**<br>**& MADRS**<br>**Reduction at End of**<br>**Open Phase** | **Serious Adverse**<br>**Events** | **Adverse Events** |
| --- | --- | --- | --- | --- | --- |
| 1<br>25-30<br>Female | Contamination<br>Onset age 10-15<br>4 antidepressants / 3 antipsychotics<br>Major depressive disorder<br>YBOCS 32 | Clomipramine 150 mg<br>Quetiapine 100 mg<br>Olanzapine 5 mg<br>DBS Right Hemisphere:<br>C+ 8-9- 4.5 V / 90 µs / 130 Hz<br>DBS Left Hemisphere:<br>C+ 0-1- 4.5 V / 90 µs / 130 Hz | YBOCS = 53.1<br>MADRS = 57.1 | Nil | Parasomnia (sleepwalking) |
| 2<br>25-30<br>Male | Harming others / Sexuality / Blasphemy<br>Onset age 5-10<br>16 antidepressants / 3 antipsychotics / ECT<br>Major depressive disorder<br>YBOCS 33 | Tranlycypromine 30 mg<br>Nortriptyline 75 mg<br>Diazepam 5mg<br>DBS Right Hemisphere:<br>C+ 9- 3.5 V / 90 µs / 130 Hz<br>DBS Left Hemisphere:<br>C+ 1- 3.5 V / 90 µs / 130 Hz | YBOCS = 69.7<br>MADRS = 70.6 | Deviation of one DBS<br>electrode during<br>implantation requiring<br>removal and re-<br>implantation. | Pustule at IPG site<br>Lead tightening behind ear<br>Reduced libido |
| 3<br>55-60<br>Male | Sexuality<br>Onset age 15-20<br>12 antidepressants / 7 antipsychotics<br>Nil<br>YBOCS 29 | Clomipramine 50 mg<br>DBS Right Hemisphere:<br>C+ 9- 4.5 V / 90 µs / 130 Hz<br>DBS Left Hemisphere:<br>C+ 1- 4.5 V / 90 µs / 130 Hz | YBOCS = 51.7<br>MADRS = 50.0 | Nil | Nil |
| 4<br>50-55<br>Male | Harming others<br>Onset age 15-20<br>9 antidepressants / 4 antipsychotics / ECT<br>Major depressive disorder<br>YBOCS 35 | Clomipramine 200 mg<br>Quetiapine XR 400 mg<br>Clonazepam 1.5 mg<br>DBS Right Hemisphere:<br>C+ 9- 10- 4.7 V / 90 µs / 130 Hz<br>DBS Left Hemisphere:<br>C+ 1- 2- 4.7 V / 90 µs / 130 Hz | YBOCS = 54.3<br>MADRS = 35.3 | Two inpatient psychiatric<br>admissions to manage<br>recurrence of depressive<br>symptoms | Nil |
| 5<br>55-60<br>Female | Sexuality / Symmetry<br>Onset age 5-10<br>9 antidepressants / 4 antipsychotics / ECT /<br>rTMS<br>Major depressive disorder / body<br>dysmorphic disorder<br>YBOCS 32 | Sertraline 100 mg<br>Pregabalin 150 mg<br>Clonazepam 0.25 mg<br>DBS Right Hemisphere:<br>C+ 9- 4.5 V / 90 µs / 130 Hz<br>DBS Left Hemisphere:<br>C+ 1- 4.5 V / 90 µs / 130 Hz | YBOCS = 28.1<br>MADRS = 46.2 | Infection of IPG requiring<br>DBS device explantation | Nil |
| 6<br>45-50<br>Female | Contamination<br>Onset aged 5-10<br>19 antidepressants / 5 antipsychotics / ECT / | Tranlycypromine 10 mg<br>Imipramine 50 mg<br>Clonazepam 0.5 mg | YBOCS = 0<br>MADRS = -4.0 | Five inpatient psychiatric<br>admissions to manage<br>persistence of obsessive & | Nil |
| | rTMS<br>Major depressive disorder<br>YBOCS 28 | Olanzapine 10 mg<br>Quetiapine IR 100 mg<br>Lithium XR 450 mg<br>DBS Right Hemisphere:<br>C+ 10- 5.6 V / 120 μs / 130 Hz<br>DBS Left Hemisphere:<br>C+ 1- 5.6 V / 120 μs / 130 Hz | | depressive symptoms | |
| 7<br>50-55<br>Male | Doubt / Perfectionism<br>Onset aged 5-10<br>3 antidepressants / 2 antipsychotics<br>Nil<br>YBOCS 35 | Clomipramine 50 mg<br>Sertraline 250 mg<br>DBS Right Hemisphere:<br>C+ 9- 10- 4.5 V / 90 μs / 130 Hz<br>DBS Left Hemisphere:<br>C+ 1- 2- 4.5 V / 90 μs / 130 Hz | YBOCS = 48.6<br>MADRS = 80.0 | Nil | Nil |
| 8<br>45-50<br>Female | Checking / Magical thinking<br>Onset aged 5-10<br>8 antidepressants / 3 antipsychotics / ECT<br>Major depressive disorder<br>YBOCS 34 | Clomipramine 125 mg<br>Desvenlafaxine 200 mg<br>Olanzapine 5mg<br>DBS Right Hemisphere:<br>C+ 9- 10- 4.5 V / 90 μs / 130 Hz<br>DBS Left Hemisphere:<br>C+ 1- 2- 4.5 V / 90 μs / 130 Hz | YBOCS = 82.3<br>MADRS = 78.9 | Nil | Nil |
| 9<br>55-60<br>Male | Checking / Doubt<br>Onset aged 5-10<br>5 antidepressants / 2 antipsychotics<br>Nil<br>YBOCS 36 | Fluoxetine 80 mg<br>Dexamphetamine 60 mg<br>DBS Right Hemisphere:<br>C+ 9- 10- 5.0 V / 90 μs / 130 Hz<br>DBS Left Hemisphere:<br>C+ 1- 2- 5.0 V / 90 μs / 130 Hz | YBOCS = 58.3<br>MADRS = 77.8 | Nil | Nil |

§ Consistent with MedRxiv stipulations, only 5-year age range is given to prevent participant identification
† For brevity, details of past psychotherapies not listed here
‡ On the quadripolar electrode, contacts are numbered 8-11 in the right hemisphere and 0-3 in the left hemisphere
Abbreviations: ECT = electroconvulsive therapy, IPG = implantable pulse generator, IR = immediate release, MADRS = Montgomery Åsberg Depression Rating Scale, rTMS = repetitive Transcranial Magnetic Stimulation, XR = extended release, YBOCS = Yale-Brown Obsessive-Compulsive Scale

**Figure 1.**
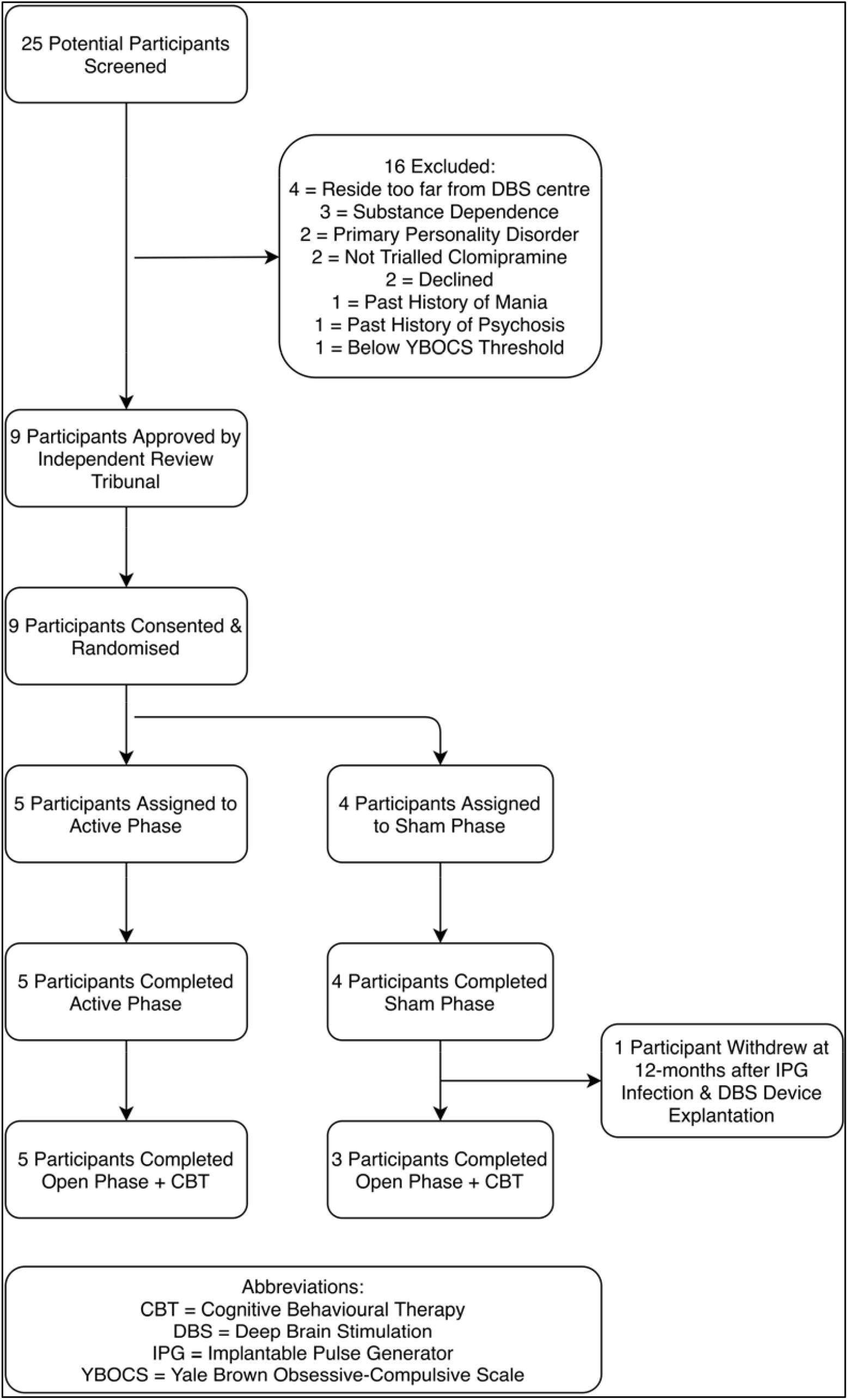
Flow Diagram of Participant Recruitment, Randomisation & Treatment.

**Figure 2.**
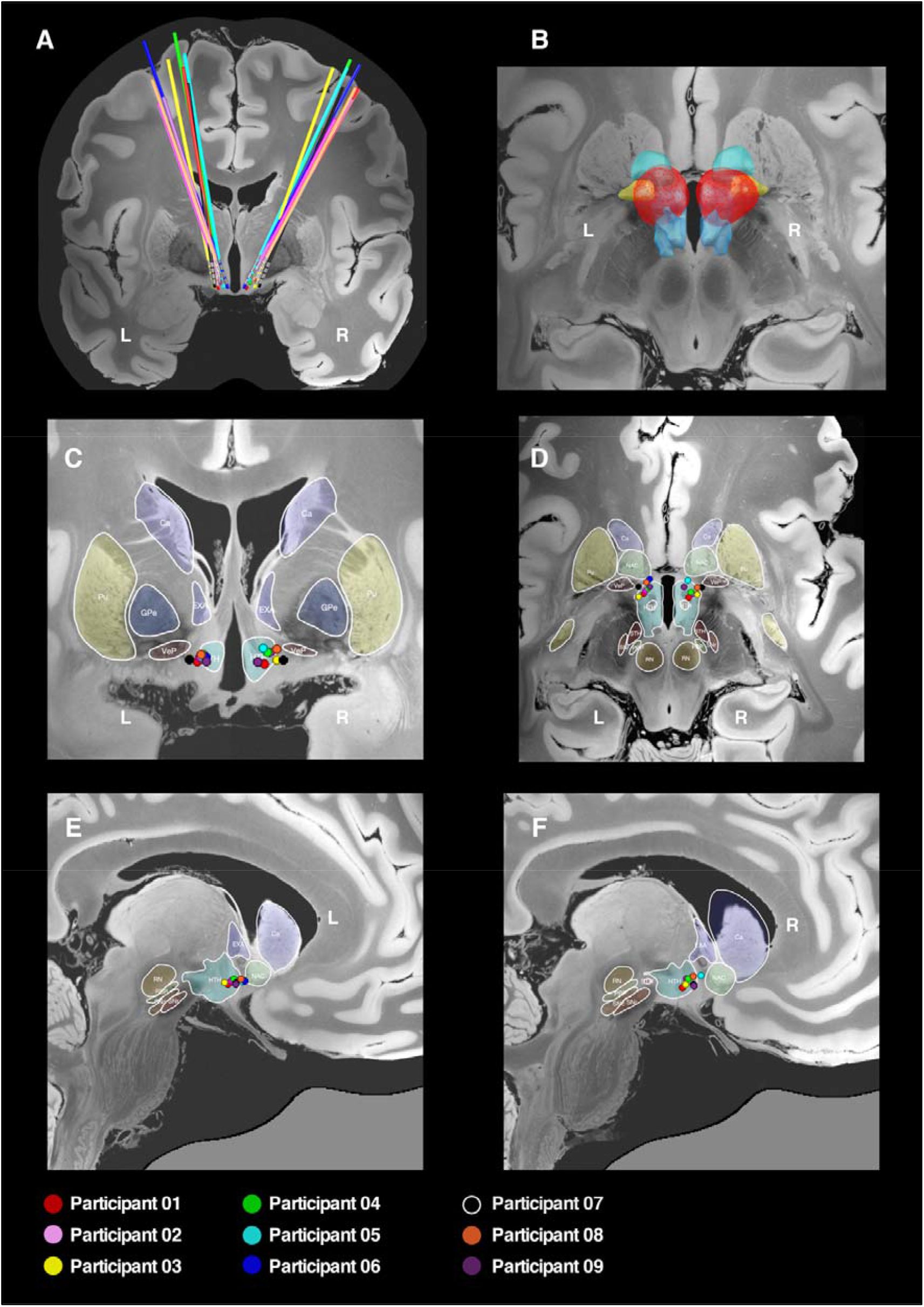
Localisation of Electrodes & Active Contacts. DBS electrodes were localised with the Lead-DBS toolbox and represented in common ICBM 2009b nonlinear asymmetric space incorporating a 7-Tesla MRI at 100 micron resolution (Edlow *et al*., 2019), with subcortical parcellations derived from a recent high-resolution atlas (Pauli *et al*., 2018). A: 3-dimensional reconstruction in the coronal plane showing electrode trajectories for the nine participants. B: 3-dimensional reconstruction in the axial plane showing the distribution of the aggregated stimulation field across the cohort (red), which can be seen to encompass the posterior segment of the nucleus accumbens (light green), the ventral pallidum (yellow) and the hypothalamus (blue). C: 2-dimensional reconstruction of active contacts in coronal plane. D: 2-dimensional reconstruction of active contacts in axial plane. E and F: 2-dimensional reconstruction of active contacts in sagittal plane. In the 2-dimensional representations coloured circles represent the second most inferior contact on each electrode (i.e. contact 9 on right electrode and contact 1 on left electrode). Abbreviations: Ca = caudate, EXA = extended amygdala (BNST), GPe = globus pallidus external segment, HTH = hypothalamus, NAC = nucleus accumbens, PBP = parabrachial pigmented nucleus, Pu = putamen, SN = substantia nigra, STH = subthalamic nucleus, RN = red nucleus, VeP = ventral pallidum

### 4.2 Outcomes

In the blinded (on versus sham) phase, there was a statistically significant difference in YBOCS reduction in favour of active stimulation (*t* = -2.9, *p* = 0.025, mean difference 4.9 points, 95 % CI = 0.8-8.9) (Figure 3). There was no significant difference in MADRS reduction (*t* = -1.1, *p* = 0.30, mean difference 3.4 points, 95 % CI = -3.7-10.5).

**Figure 3.**
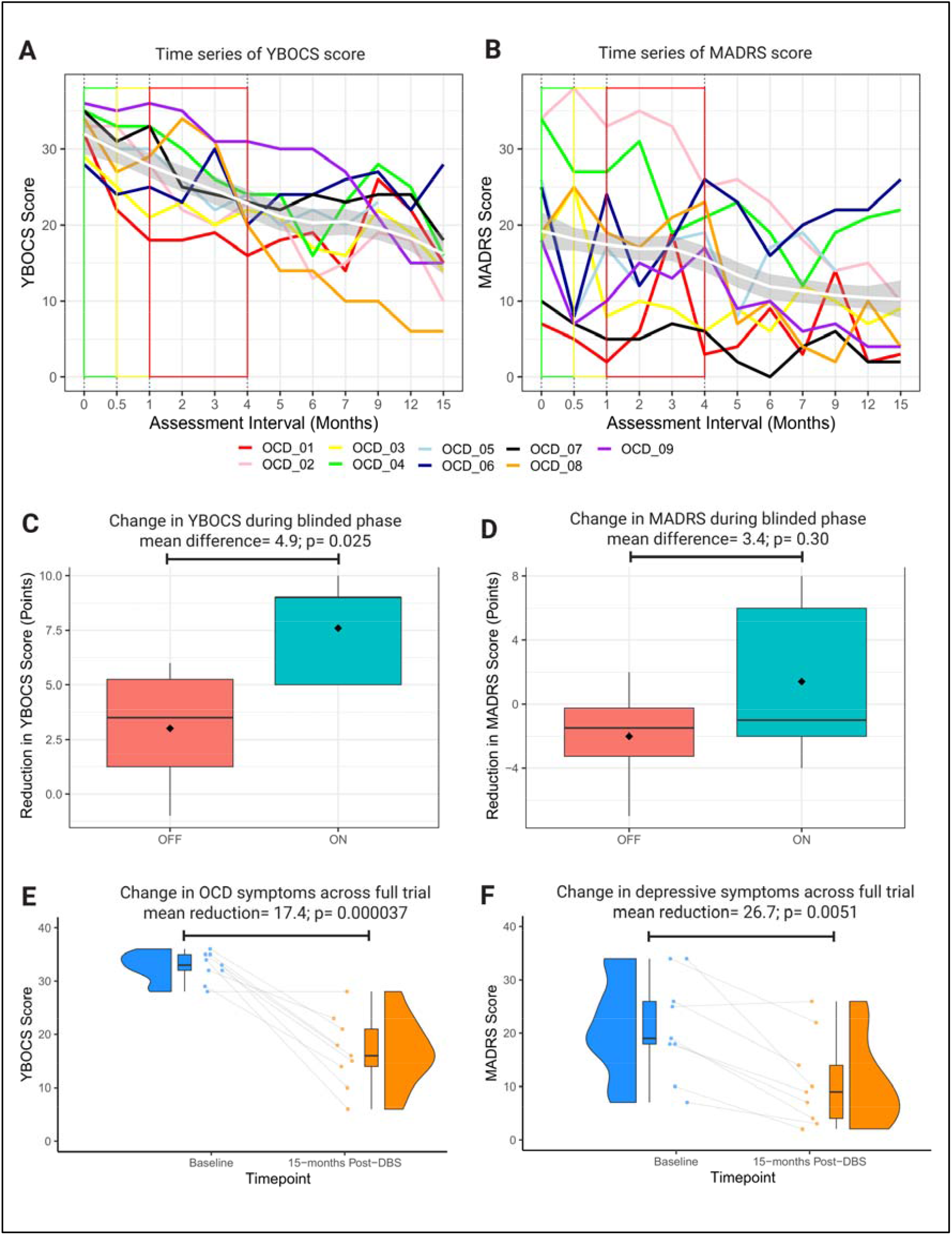
Participant Outcomes. A and B: Time series of individual participant outcomes for primary (YBOCS) and secondary (MADRS) variables. Within each graph, group average trajectory is represented by a loess smoothed curve (white) ± 1 standard error (grey). Baseline measurement denoted by green outline, recovery phase by yellow outline and blinded phase by red outline. C and D: Boxplots of YBOCS and MADRS change by randomised group (on = green versus off = red) during the blinded phase. E and F: Raincloud plots of YBOCS and MADRS change across the full trial. Raincloud plots made with code provided by Allen *et al*. (2019) and van Langen. (2020).

After one year of open-label stimulation and a course of ERP-based CBT, the mean reduction in YBOCS was 17.4 ±2.0 points (*Χ*^2^ (11) = 39.9, p = 3.7 × 10^−5^) with no statistically significant covariates (Figure 3). Seven participants were responders as defined by the 35 % YBOCS reduction criterion, with a mean percentage reduction across the cohort of 49.6 ±23.7. ERP-based CBT commenced an average of 10.1 ±2.6 months after DBS with a mean additive YBOCS reduction of 4.8 ±3.9 points (*t* = -3.5, *p* = 0.011, 95 % CI = 1.5-8.0). The mean reduction in MADRS was 10.8 ±2.5 points (*Χ*^2^ (11) = 26.7, p = 0.0051) with age (*t* = -2.7, *p* = 0.0084) and baseline MADRS (*t* = 13.4, *p* = 2.0 × 10^−16^) being significant covariates. Six participants were responders as defined by the 50 % MADRS reduction criterion, with a mean percentage reduction across the cohort of 54.7 ±27.2.

### 4.3 Relationship of Structural Connectivity to YBOCS Reduction

The local dispersion of the stimulation field within neighbouring subcortical structures including the NAcc, ventral pallidum, hypothalamus and terminal fibres of the stria terminalis was not related to relief of OCD symptoms (Supplementary Material). Using a normative connectome to identify white matter fibres connected to the stimulation field in each hemisphere for each participant, those connections most highly associated with YBOCS reduction were found in the right hemisphere (Figure 4A). These included a tract passing through the midbrain, traversing the BNST and onwards to the right ventrolateral prefrontal cortex. A tract connecting the BNST with the right amygdala was also identified, with connecting fibres passing through the hippocampal white matter and traversing back into the BNST via the fornix.

**Figure 4.**
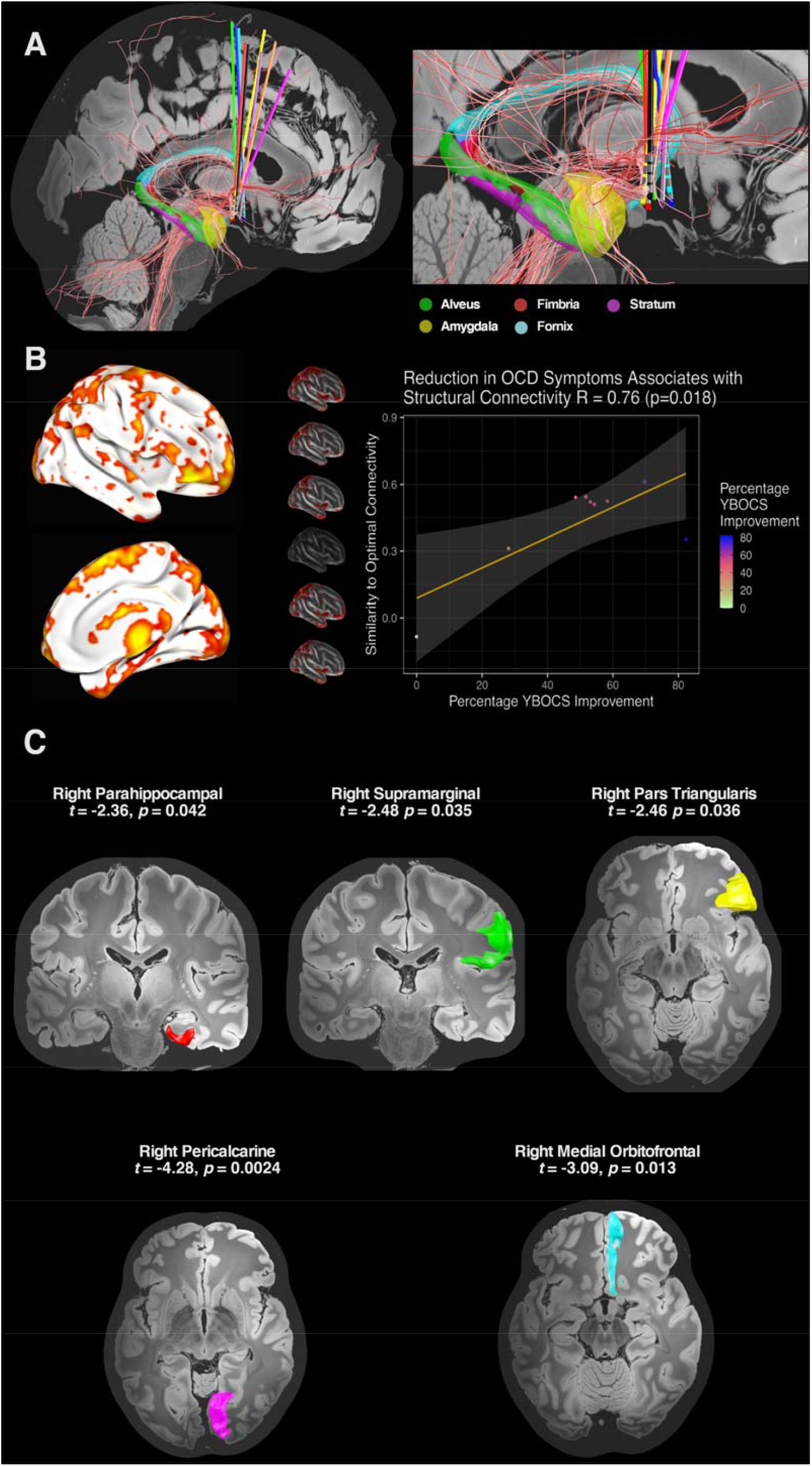
Structural Connectivity & YBOCS Reduction. A: White matter fibres connected to the stimulation field and discriminative of outcome were isolated in the right hemisphere. These included a fibre tract passing through the midbrain to the ventrolateral prefrontal cortex and a fibre tract connecting the site of stimulation with the amygdala. Fibres in this region also passed through the hippocampal white matter and returned to the BNST via the stria terminalis adjacent to the fornix. Subcortical parcellations of the amygdala, hippocampus and fornix were derived from recent automated segmentation methods (Amaral *et al*., 2018; Entis *et al*., 2012; Pipitone *et al*., 2014). B: An optimal connectivity profile was generated by identifying those brain voxels structurally connected with the stimulation field and most highly correlated with YBOCS reduction. Cortical regions implicated in this optimal right-hemispheric ‘R-map’ included ventromedial and ventrolateral prefrontal cortex, dorsomedial prefrontal cortex, medial temporal cortex, parietal cortex and extrastriate visual cortex. These findings were corroborated in a leave-one-out cross-validation, in which each participant’s percentage YBOCS reduction was predicted by comparing their structural connectivity profile with an optimal connectivity map derived from the remaining participants. C: In a region of interest analysis, cortical regions derived from the R-map were tested in a multivariate linear mixed-effects model for their association with YBOCS reduction.

An ‘optimal’ connectivity map derived from correlating each brain voxel (weighted by structural connectivity) to YBOCS reduction also identified the right ventrolateral and parahippocampal regions, as well as right extrastriate, parietal and dorsomedial prefrontal areas (Figure 4B and local maxima Supplementary Table 1). In a leave-one-out cross-validation, structural connectivity of the stimulation field was significantly associated with YBOCS reduction (*r* = 0.76, *p* = 0.018).

Based on these findings, corresponding cortical regions derived from the Desikan-Killiany-Tourville labelling protocol were entered into the multivariate, linear mixed-effects model. Structural connectivity of the right hemispheric stimulation field with right orbitofrontal (*t* = -3.1, *p* = 0.013), right parahippocampal (*t* = -2.4, *p* = 0.042), right pars triangularis (*t* = -2.5, *p* = 0.036), right pericalcarine (*t* = -4.3, *p* = 0.0024) and right supramarginal regions (*t* = -2.5, *p* = 0.035) was significantly associated with YBOCS reduction. Connectivity with the right paracentral (*t* = -1.8, *p* = 0.11) region was not statistically significant. Univariate correlations displayed in Supplementary Material.

### 4.4 Adverse Events

There were nine serious adverse events (SAEs) affecting four participants (Table 1). Five of these were attributable to one participant (06) who was a non-responder and was readmitted to hospital to manage persistent psychiatric symptoms. A further participant (04) was readmitted to hospital on two occasions to manage a recurrence of depressive symptoms.

Two SAEs were device-related. One participant (02) required re-siting of a DBS electrode that had migrated 3 mm from the target during implantation. This was accomplished without any further complication. One participant (05) developed an infection of the IPG that migrated to the extension leads necessitating removal of the DBS device. There were four adverse events affecting two participants (Table 1). These were transient in nature except for reduced libido (participant 02), which persisted throughout follow up. Notably, there were no psychiatric adverse effects considered to be device related. All participants (except 05 who withdrew) required IPG replacement due to battery depletion during the study.

## 5 DISCUSSION

In nine participants with severe, treatment-refractory OCD, we demonstrate that DBS of the BNST substantially alleviated symptoms, with a mean YBOCS reduction of 49.6 % and seven participants meeting the threshold for clinically-significant response after 12-months of open-label stimulation. Moreover, we describe a statistically-significant benefit of active stimulation over sham during a 3-month, double-blind, delayed-onset phase. Our data adds to the emerging literature supporting the use of DBS as a therapy in otherwise treatment-resistant OCD and specifically reproduces prior work targeting the BNST. (Luyten *et al*., 2016) Open-label stimulation also significantly reduced co-morbid depressive symptoms, although this result should be viewed with more circumspection as depression was not a primary target of the intervention and two participants reported only mild symptoms at baseline.

Extending prior clinical findings, we also characterise a subcortical structural connectivity profile associated with optimal response to DBS at this target. Here, a right-hemispheric tract traversing the stimulation field and associated with YBOCS reduction connected the BNST to the amygdala. Connected fibres also involved the hippocampal formation and fornix, which form part of the circuit of Papez (Papez, 1995). From a physiological perspective, the BNST functions as a component of the ‘extended amygdala’ and drives a state of sustained apprehension (anxiety) (Lebow and Chen, 2016). Of note, recent work has also demonstrated a central role for the amygdala in mediating a rapid reduction in anxiety symptoms after ALIC DBS for OCD (Fridgeirsson *et al*., 2020), which heralds later improvement in obsessions and compulsions. Overall, this supports the role of DBS in facilitating fear extinction through reducing anxiety. Aberrant fear conditioning (enhanced acquisition and impaired extinction) is a central construct in the development and maintenance of OCD (Geller *et al*., 2017; Milad *et al*., 2013), and may explain why more severely affected individuals cannot tolerate or do not respond to exposure-oriented CBT (Geller *et al*., 2019). This may also explain why, after DBS, our participants were now able to tolerate, and accrue a statistically-significant additional benefit from CBT during open stimulation, consistent with previous work (Mantione *et al*., 2014).

Importantly, this improvement in fear extinction may be mediated via enhanced top-down input to the amygdala from the prefrontal cortex (Fridgeirsson *et al*., 2020). In our cohort, fibres associated with YBOCS reduction were also characterised passing to the prefrontal cortex and potentially representing a structural correlate of this effect. This connectivity profile was similar in distribution to that previously described by other centres employing different targets such as the NAcc / ALIC interface and the STN (Baldermann *et al*., 2019; Li *et al*., 2019). These findings support the existence of a common anatomical substrate that underpins response across discrete sites, as well as being consistent with prior work demonstrating that alterations in frontostriatal connectivity are implicated in the response to NAcc / ALIC DBS (Figee *et al*., 2011). Moreover, the distribution of connected fibres associated with YBOCS reduction was strikingly similar to prior research characterising the structural connectivity of the BNST in healthy participants (Kruger *et al*., 2015).

Connectivity of the stimulation field with right-hemispheric cortical regions of interest in the prefrontal, temporal, parietal and occipital lobes was also significantly associated with YBOCS reduction. Interestingly, these same regions have previously been implicated in morphometric analyses of structural connectivity, grey matter volume, cortical thickness, surface area and gyrification amongst individuals with OCD (Fan *et al*., 2013; Peng *et al*., 2014; Rotge *et al*., 2010; Yun *et al*., 2020), suggesting that there may be a neuroanatomic ‘fingerprint’ of susceptibility to OCD that is modulated by DBS. Importantly, using cross-validation, YBOCS reduction could be accurately predicted in a single participant by comparing their connectivity profile to a pooled analysis of the connectivity amongst the remainder of the cohort. This suggests that the recruitment of specific fibre pathways by the stimulation field is an important determinant of outcome. More generally, the right lateralisation of our findings is interesting given previous work that implicates right-hemispheric corticostriatal circuits in inhibition (Aron *et al*., 2004; Aron *et al*., 2007; Rae *et al*., 2015), impulsivity after subthalamic DBS for Parkinson’s disease (Mosley *et al*., 2018; Mosley *et al*., 2020b), and reduction of OCD symptoms after NAcc / ALIC DBS (Baldermann *et al*., 2019).

Serious adverse events were predominantly accounted for by persisting psychiatric symptoms in a non-responder with repeated readmissions to hospital. It is noteworthy that the connectivity profile of this individual was most distinct from the rest of the cohort with electrodes that were more anteromedial and a stimulation field that was less connected to the right-hemispheric regions of interest (Figures 2 and 4, Supplementary Figure 2), suggesting a potential explanation for this lack of response. IPG infection and device removal was the most significant device-related event, affecting one participant. No participants developed stimulation-related psychiatric side effects such as agitation, impulsivity and hypomania, as has previously been reported (Denys *et al*., 2010; Greenberg *et al*., 2010; Tyagi *et al*., 2019). This may have been attributable to our deliberately slow titration protocol and the use of lower stimulation amplitudes than have previously been described. However, despite the use of more modest amplitudes, IPG depletion occurred in all participants before the close of the trial, necessitating replacement.

The use of a staggered-onset rather than a crossover design in the double-blind phase could be considered a limitation. In previous trials using a crossover design (Denys *et al*., 2010; Luyten *et al*., 2016; Mallet *et al*., 2008), optimal stimulation settings were already determined after an open-phase, increasing the likelihood of a treatment effect in the active condition. However, based on prior work describing a significant rebound of aversive OCD symptoms after therapy interruption (Ooms *et al*., 2014), we considered it more ethically acceptable to delay treatment rather than cease a treatment that had previously been effective. Moreover, one significant benefit of our approach was that the likelihood of participants becoming unblinded by sensations associated with active stimulation was minimised.

The use of normative rather than participant-specific connectivity data is a further limitation and has been discussed elsewhere (Coenen *et al*., 2019; Li *et al*., 2019; Treu *et al*., 2020). Whilst participant-specific anatomical variability is lost, the quality of these group-average datasets is high and curated by teams with longstanding expertise. The reliability of analyses derived from these data may therefore be acceptable, and normative connectomic data has been employed to make out-of-sample predictions across disorders and treatment modalities (Al-Fatly *et al*., 2019; Baldermann *et al*., 2019; Horn *et al*., 2017; Joutsa *et al*., 2018; Weigand *et al*., 2018). Thus, whilst normative data should not be the basis for surgical decision-making in one individual, it may yield important insights into mechanisms of disease and treatment-response within and across cohorts.

In summary, in a cohort of participants with severe, treatment-refractory OCD, we demonstrate that active stimulation at the BNST is superior to placebo in a randomised, double-blind, sham-controlled, delayed-onset clinical trial, with a further significant benefit accrued following a longer phase of open-label stimulation incorporating a course of ERP-based CBT. We also delineate a structural connectivity profile associated with clinical response, which comprised subcortical regions implicated in fear conditioning and emotional processing, as well as cortical regions implicated in prior morphometric analyses of persons with OCD. We anticipate that our findings will motivate more precise targeting of stimulation within these networks, using participant-specific connectivity data to optimise treatment at the individual level, as has been described in DBS for treatment-resistant major depression (Riva-Posse *et al*., 2014; Riva-Posse *et al*., 2018).

## 6 ACKNOWLEDGEMENTS

The authors gratefully acknowledge the commitment of participants who contributed their time to this study. The authors acknowledge the support of St Andrew’s War Memorial Hospital and the Queensland Mental Health Review Tribunal. The authors thank Dr Greg Apel, Ms Lisa McKeown, Ms Sara Gottliebsen and Ms Brenda Rosser for their administrative contributions to the development and operation of the trial. The authors also thank the data monitoring and safety board members Prof Michael Breakspear, Dr Jim Rodney and Dr Josh Geffen. Finally, the authors thank Dr Andreas Horn and Dr Ningfei Li from the Lead-DBS development team for technical assistance with the modelling of fibre tracts.

## 7 FUNDING AGENCIES

The trial was funded by the University of Queensland through the Queensland Brain Institute in partnership with Medtronic. Medtronic provided the PC+S devices, 3389 stimulating electrodes, physician programmers and related equipment. Medtronic also made a cash contribution to the research costs. Medtronic had no role in the design of the experimental protocol, data collection, analysis or writing of the manuscript.

## 9 ETHICS APPROVAL

Prior to the commencement of data collection, the full protocol was approved by the Human Research Ethics Committees of the University of Queensland and UnitingCare Health. All participants gave written, informed consent to participate in the study. All procedures were carried out in accordance with the approved protocol.

## REFERENCES

Abelson, J.L., Curtis, G.C., Sagher, O., Albucher, R.C., Harrigan, M., Taylor, S.F., et al., 2005. Deep brain stimulation for refractory obsessive-compulsive disorder. Biol Psychiatry. 57, 510–6.

Accolla, E.A., Herrojo Ruiz, M., Horn, A., Schneider, G.H., Schmitz-Hubsch, T., Draganski, B., et al., 2016. Brain networks modulated by subthalamic nucleus deep brain stimulation. Brain. 139, 2503–15.

Akram, H., Sotiropoulos, S.N., Jbabdi, S., Georgiev, D., Mahlknecht, P., Hyam, J., et al., 2017. Subthalamic deep brain stimulation sweet spots and hyperdirect cortical connectivity in Parkinson’s disease. Neuroimage. 158, 332–345.

Al-Fatly, B., Ewert, S., Kubler, D., Kroneberg, D., Horn, A., Kuhn, A.A., 2019. Connectivity profile of thalamic deep brain stimulation to effectively treat essential tremor. Brain. 142, 3086–3098.

Allen, M., Poggiali, D., Whitaker, K., Marshall, T., Kievit, R., 2019. Raincloud plots: a multi-platform tool for robust data visualization [version 1; peer review: 2 approved]. Wellcome Open Research.4.

Amaral, R.S.C., Park, M.T.M., Devenyi, G.A., Lynn, V., Pipitone, J., Winterburn, J., et al., 2018. Manual segmentation of the fornix, fimbria, and alveus on high-resolution 3T MRI: Application via fully-automated mapping of the human memory circuit white and grey matter in healthy and pathological aging. Neuroimage. 170, 132–150.

American Psychiatric Association., 2013. Diagnostic and statistical manual of mental disorders: DSM-5, Vol., American Psychiatric Publishing, Washington, DC.

Aron, A.R., Robbins, T.W., Poldrack, R.A., 2004. Inhibition and the right inferior frontal cortex. Trends Cogn Sci. 8, 170–7.

Aron, A.R., Behrens, T.E., Smith, S., Frank, M.J., Poldrack, R.A., 2007. Triangulating a cognitive control network using diffusion-weighted magnetic resonance imaging (MRI) and functional MRI. J Neurosci. 27, 3743–52.

Avants, B.B., Epstein, C.L., Grossman, M., Gee, J.C., 2008. Symmetric diffeomorphic image registration with cross-correlation: evaluating automated labeling of elderly and neurodegenerative brain. Med Image Anal. 12, 26–41.

Baldermann, J.C., Melzer, C., Zapf, A., Kohl, S., Timmermann, L., Tittgemeyer, M., et al., 2019. Connectivity Profile Predictive of Effective Deep Brain Stimulation in Obsessive-Compulsive Disorder. Biol Psychiatry. 85, 735–743.

Benabid, A.L., Pollak, P., Gross, C., Hoffmann, D., Benazzouz, A., Gao, D.M., et al., 1994. Acute and long-term effects of subthalamic nucleus stimulation in Parkinson’s disease. Stereotact Funct Neurosurg. 62, 76–84.

Bloch, M.H., Landeros-Weisenberger, A., Kelmendi, B., Coric, V., Bracken, M.B., Leckman, J.F., 2006. A systematic review: antipsychotic augmentation with treatment refractory obsessive-compulsive disorder. Mol Psychiatry. 11, 622–32.

Chen, Y., Ge, S., Li, Y., Li, N., Wang, J., Wang, X., et al., 2018. Role of the Cortico-Subthalamic Hyperdirect Pathway in Deep Brain Stimulation for the Treatment of Parkinson Disease: A Diffusion Tensor Imaging Study. World Neurosurg. 114, e1079–e1085.

Coenen, V.A., Schlaepfer, T.E., Varkuti, B., Schuurman, P.R., Reinacher, P.C., Voges, J., et al., 2019. Surgical decision making for deep brain stimulation should not be based on aggregated normative data mining. Brain Stimul. 12, 1345–1348.

Denys, D., Mantione, M., Figee, M., van den Munckhof, P., Koerselman, F., Westenberg, H., et al., 2010. Deep brain stimulation of the nucleus accumbens for treatment-refractory obsessive-compulsive disorder. Arch Gen Psychiatry. 67, 1061–8.

Dougherty, D.D., 2019. Will Deep Brain Stimulation Help Move Precision Medicine to the Clinic in Psychiatry? Biol Psychiatry. 85, 706–707.

Edlow, B.L., Mareyam, A., Horn, A., Polimeni, J.R., Witzel, T., Tisdall, M.D., et al., 2019. 7 Tesla MRI of the ex vivo human brain at 100 micron resolution. Sci Data. 6, 244.

Entis, J.J., Doerga, P., Barrett, L.F., Dickerson, B.C., 2012. A reliable protocol for the manual segmentation of the human amygdala and its subregions using ultra-high resolution MRI. Neuroimage. 60, 1226–35.

Erzegovesi, S., Cavallini, M.C., Cavedini, P., Diaferia, G., Locatelli, M., Bellodi, L., 2001. Clinical predictors of drug response in obsessive-compulsive disorder. J Clin Psychopharmacol. 21, 488–92.

Fan, Q., Palaniyappan, L., Tan, L., Wang, J., Wang, X., Li, C., et al., 2013. Surface anatomical profile of the cerebral cortex in obsessive-compulsive disorder: a study of cortical thickness, folding and surface area. Psychol Med. 43, 1081–91.

Farrand, S., Evans, A.H., Mangelsdorf, S., Loi, S.M., Mocellin, R., Borham, A., et al., 2018. Deep brain stimulation for severe treatment-resistant obsessive-compulsive disorder: An open-label case series. Aust N Z J Psychiatry. 52, 699–708.

Figee, M., Vink, M., de Geus, F., Vulink, N., Veltman, D.J., Westenberg, H., et al., 2011. Dysfunctional reward circuitry in obsessive-compulsive disorder. Biol Psychiatry. 69, 867–74.

Fridgeirsson, E.A., Figee, M., Luigjes, J., van den Munckhof, P., Schuurman, P.R., van Wingen, G., et al., 2020. Deep brain stimulation modulates directional limbic connectivity in obsessive-compulsive disorder. Brain. 143, 1603–1612.

Geller, D.A., McGuire, J.F., Orr, S.P., Pine, D.S., Britton, J.C., Small, B.J., et al., 2017. Fear conditioning and extinction in pediatric obsessive-compulsive disorder. Ann Clin Psychiatry. 29, 17–26.

Geller, D.A., McGuire, J.F., Orr, S.P., Small, B.J., Murphy, T.K., Trainor, K., et al., 2019. Fear extinction learning as a predictor of response to cognitive behavioral therapy for pediatric obsessive compulsive disorder. J Anxiety Disord. 64, 1–8.

Goodman, W.K., Price, L.H., Rasmussen, S.A., Mazure, C., Fleischmann, R.L., Hill, C.L., et al., 1989. The Yale-Brown Obsessive Compulsive Scale. I. Development, use, and reliability. Arch Gen Psychiatry. 46, 1006–11.

Goodman, W.K., Foote, K.D., Greenberg, B.D., Ricciuti, N., Bauer, R., Ward, H., et al., 2010. Deep brain stimulation for intractable obsessive compulsive disorder: pilot study using a blinded, staggered-onset design. Biol Psychiatry. 67, 535–42.

Greenberg, B.D., Malone, D.A., Friehs, G.M., Rezai, A.R., Kubu, C.S., Malloy, P.F., et al., 2006. Three-year outcomes in deep brain stimulation for highly resistant obsessive-compulsive disorder. Neuropsychopharmacology. 31, 2384–93.

Greenberg, B.D., Gabriels, L.A., Malone, D.A., Jr., Rezai, A.R., Friehs, G.M., Okun, M.S., et al., 2010. Deep brain stimulation of the ventral internal capsule/ventral striatum for obsessive-compulsive disorder: worldwide experience. Mol Psychiatry. 15, 64–79.

Horn, A., Kuhn, A.A., 2015. Lead-DBS: a toolbox for deep brain stimulation electrode localizations and visualizations. Neuroimage. 107, 127–35.

Horn, A., Reich, M., Vorwerk, J., Li, N., Wenzel, G., Fang, Q., et al., 2017. Connectivity Predicts deep brain stimulation outcome in Parkinson disease. Ann Neurol. 82, 67–78.

Horn, A., Li, N., Dembek, T.A., Kappel, A., Boulay, C., Ewert, S., et al., 2019. Lead-DBS v2: Towards a comprehensive pipeline for deep brain stimulation imaging. Neuroimage. 184, 293–316.

Issakidis, C., Andrews, G., 2002. Service utilisation for anxiety in an Australian community sample. Soc Psychiatry Psychiatr Epidemiol. 37, 153–63.

Joutsa, J., Horn, A., Hsu, J., Fox, M.D., 2018. Localizing parkinsonism based on focal brain lesions. Brain. 141, 2445–2456.

Kessler, R.C., Berglund, P., Demler, O., Jin, R., Merikangas, K.R., Walters, E.E., 2005. Lifetime prevalence and age-of-onset distributions of DSM-IV disorders in the National Comorbidity Survey Replication. Arch Gen Psychiatry. 62, 593–602.

Klein, A., Tourville, J., 2012. 101 labeled brain images and a consistent human cortical labeling protocol. Front Neurosci. 6, 171.

Klein, A., Ghosh, S.S., Bao, F.S., Giard, J., Hame, Y., Stavsky, E., et al., 2017. Mindboggling morphometry of human brains. PLoS Comput Biol. 13, e1005350.

Kruger, O., Shiozawa, T., Kreifelts, B., Scheffler, K., Ethofer, T., 2015. Three distinct fiber pathways of the bed nucleus of the stria terminalis to the amygdala and prefrontal cortex. Cortex. 66, 60–8.

Kuznetsova, A., Brockhoff, P.B., Christensen, R.H.B., 2017. lmerTest Package: Tests in Linear Mixed Effects Models. 2017. 82, 26.

Lebow, M.A., Chen, A., 2016. Overshadowed by the amygdala: the bed nucleus of the stria terminalis emerges as key to psychiatric disorders. Mol Psychiatry. 21, 450–63.

Li, N., Baldermann, J.C., Kibleur, A., Treu, S., Elias, G., Boutet, A., et al., 2019. Toward a unified connectomic target for deep brain stimulation in obsessive-compulsive disorder, Vol., 10.1101/608786.

Luyten, L., Hendrickx, S., Raymaekers, S., Gabriels, L., Nuttin, B., 2016. Electrical stimulation in the bed nucleus of the stria terminalis alleviates severe obsessive-compulsive disorder. Mol Psychiatry. 21, 1272–80.

Mallet, L., Polosan, M., Jaafari, N., Baup, N., Welter, M.L., Fontaine, D., et al., 2008. Subthalamic nucleus stimulation in severe obsessive-compulsive disorder. N Engl J Med. 359, 2121–34.

Mantione, M., Nieman, D.H., Figee, M., Denys, D., 2014. Cognitive-behavioural therapy augments the effects of deep brain stimulation in obsessive-compulsive disorder. Psychol Med. 10.1017/S0033291714000956, 1-8.

Mathers, C., Fat, D.M., Boerma, J.T., 2008. The global burden of disease 2004 update, Vol., World Health Organization, Geneva, Switzerland.

Milad, M.R., Furtak, S.C., Greenberg, J.L., Keshaviah, A., Im, J.J., Falkenstein, M.J., et al., 2013. Deficits in conditioned fear extinction in obsessive-compulsive disorder and neurobiological changes in the fear circuit. JAMA Psychiatry. 70, 608-18; quiz 554.

Montgomery, S.A., Asberg, M., 1979. A new depression scale designed to be sensitive to change. Br J Psychiatry. 134, 382–9.

Mosley, P., Paliwal, S., Robinson, K., Coyne, T., Silburn, P., Tittgemeyer, M., et al., 2020a. The Structural Connectivity of Subthalamic Deep Brain Stimulation Correlates with Impulsivity in Parkinson’s. Brain. 10.1093/brain/awaa148, In Press.

Mosley, P.E., Smith, D., Coyne, T., Silburn, P., Breakspear, M., Perry, A., 2018. The site of stimulation moderates neuropsychiatric symptoms after subthalamic deep brain stimulation for Parkinson’s disease. NeuroImage: Clinical. 18, 996–1006.

Mosley, P.E., Paliwal, S., Robinson, K., Coyne, T., Silburn, P., Tittgemeyer, M., et al., 2020b. The structural connectivity of subthalamic deep brain stimulation correlates with impulsivity in Parkinson’s. Brain. 10.1093/brain/awaa148, In Press.

Nuttin, B., Cosyns, P., Demeulemeester, H., Gybels, J., Meyerson, B., 1999. Electrical stimulation in anterior limbs of internal capsules in patients with obsessive-compulsive disorder. Lancet. 354, 1526.

Nuttin, B.J., Gabriels, L.A., Cosyns, P.R., Meyerson, B.A., Andreewitch, S., Sunaert, S.G., et al., 2003. Long-term electrical capsular stimulation in patients with obsessive-compulsive disorder. Neurosurgery. 52, 1263-72; discussion 1272-4.

Ooms, P., Blankers, M., Figee, M., Mantione, M., van den Munckhof, P., Schuurman, P.R., et al., 2014. Rebound of affective symptoms following acute cessation of deep brain stimulation in obsessive-compulsive disorder. Brain Stimul. 7, 727–31.

Papez, J.W., 1995. A proposed mechanism of emotion. 1937. J Neuropsychiatry Clin Neurosci. 7, 103–12.

Pauli, W.M., Nili, A.N., Tyszka, J.M., 2018. A high-resolution probabilistic in vivo atlas of human subcortical brain nuclei. Sci Data. 5, 180063.

Peng, Z., Shi, F., Shi, C., Yang, Q., Chan, R.C., Shen, D., 2014. Disrupted cortical network as a vulnerability marker for obsessive-compulsive disorder. Brain Struct Funct. 219, 1801–12.

Pipitone, J., Park, M.T., Winterburn, J., Lett, T.A., Lerch, J.P., Pruessner, J.C., et al., 2014. Multi-atlas segmentation of the whole hippocampus and subfields using multiple automatically generated templates. Neuroimage. 101, 494–512.

R Core Team, 2014. R: A Language and Environment for Statistical Computing. Vol., ed.^eds. R Foundation for Statistical Computing, Vienna, Austria.

Rae, C.L., Hughes, L.E., Anderson, M.C., Rowe, J.B., 2015. The prefrontal cortex achieves inhibitory control by facilitating subcortical motor pathway connectivity. J Neurosci. 35, 786–94.

Riva-Posse, P., Choi, K.S., Holtzheimer, P.E., McIntyre, C.C., Gross, R.E., Chaturvedi, A., et al., 2014. Defining Critical White Matter Pathways Mediating Successful Subcallosal Cingulate Deep Brain Stimulation for Treatment-Resistant Depression. Biol Psychiatry. 10.1016/j.biopsych.2014.03.029.

Riva-Posse, P., Choi, K.S., Holtzheimer, P.E., Crowell, A.L., Garlow, S.J., Rajendra, J.K., et al., 2018. A connectomic approach for subcallosal cingulate deep brain stimulation surgery: prospective targeting in treatment-resistant depression. Mol Psychiatry. 23, 843–849.

Rotge, J.Y., Langbour, N., Guehl, D., Bioulac, B., Jaafari, N., Allard, M., et al., 2010. Gray matter alterations in obsessive-compulsive disorder: an anatomic likelihood estimation meta-analysis. Neuropsychopharmacology. 35, 686–91.

Schuepbach, W.M., Rau, J., Knudsen, K., Volkmann, J., Krack, P., Timmermann, L., et al., 2013. Neurostimulation for Parkinson’s disease with early motor complications. N Engl J Med. 368, 610–22.

Treu, S., Strange, B., Oxenford, S., Neumann, W.J., Kuhn, A., Li, N., et al., 2020. Deep Brain Stimulation: Imaging on a group level. Neuroimage. 10.1016/j.neuroimage.2020.117018,117018.

Tyagi, H., Apergis-Schoute, A.M., Akram, H., Foltynie, T., Limousin, P., Drummond, L.M., et al., 2019. A Randomized Trial Directly Comparing Ventral Capsule and Anteromedial Subthalamic Nucleus Stimulation in Obsessive-Compulsive Disorder: Clinical and Imaging Evidence for Dissociable Effects. Biol Psychiatry. 85, 726–734.

Van Essen, D.C., Smith, S.M., Barch, D.M., Behrens, T.E., Yacoub, E., Ugurbil, K., et al., 2013. The WU-Minn Human Connectome Project: an overview. Neuroimage. 80, 62–79.

van Langen, J., 2020. Open-visualizations in R and Python (Version v.1.0.4). Vol., http://doi.org/10.5281/zenodo.3715576, ed.^eds.

Vanegas-Arroyave, N., Lauro, P.M., Huang, L., Hallett, M., Horovitz, S.G., Zaghloul, K.A., et al., 2016. Tractography patterns of subthalamic nucleus deep brain stimulation. Brain. 139, 1200–10.

Vorwerk, J., Oostenveld, R., Piastra, M.C., Magyari, L., Wolters, C.H., 2018. The FieldTrip-SimBio pipeline for EEG forward solutions. Biomed Eng Online. 17, 37.

Weigand, A., Horn, A., Caballero, R., Cooke, D., Stern, A.P., Taylor, S.F., et al., 2018. Prospective Validation That Subgenual Connectivity Predicts Antidepressant Efficacy of Transcranial Magnetic Stimulation Sites. Biol Psychiatry. 84, 28–37.

Williams, J.B., Kobak, K.A., 2008. Development and reliability of a structured interview guide for the Montgomery Asberg Depression Rating Scale (SIGMA). Br J Psychiatry. 192, 52–8.

Yun, J.Y., Boedhoe, P.S.W., Vriend, C., Jahanshad, N., Abe, Y., Ameis, S.H., et al., 2020. Brain structural covariance networks in obsessive-compulsive disorder: a graph analysis from the ENIGMA Consortium. Brain. 143, 684–700.

