## Supplementary Information for "A Randomised, Double-Blind, Sham-Controlled Trial of Deep Brain Stimulation of the Bed Nucleus of the Stria Terminalis for Treatment-Resistant Obsessive-Compulsive Disorder"

Philip E. Mosley FRANZCP PhD ^1,2,3,4^, François Windels PhD ^3^, John Morris PhD ^3^, Terry Coyne FRACS ^3,5^, Rodney Marsh FRANZCP ^2,4^, Andrea Giorni PhD ^3^, Adith Mohan MRCPsych FRANZCP ^6,7^, Perminder Sachdev FRANZCP PhD ^6,7^, Emily O’Leary PhD ^8^, Mark Boschen PhD ^9^, Pankaj Sah PhD ^3,10^ †, Peter A. Silburn FRACP PhD ^2,3^ †

*^1^ Systems Neuroscience Group, QIMR Berghofer Medical Research Institute, Herston, Queensland, Australia*

*^2^ Neurosciences Queensland, St Andrew’s War Memorial Hospital, Spring Hill, Queensland, Australia*

*^3^ Queensland Brain Institute, University of Queensland, St Lucia, Queensland, Australia*

*^4^ Faculty of Medicine, University of Queensland, Herston, Queensland, Australia*

*^5^ Brizbrain and Spine, the Wesley Hospital, Auchenflower, Queensland, Australia*

*^6^ Centre for Healthy Brain Ageing (CHeBA), School of Psychiatry, University of New South Wales, Sydney, Australia*

*^7^ Neuropsychiatric Institute, The Prince of Wales Hospital, Randwick, New South Wales, Australia.*

*^8^ The OCD Clinic, Bulimba, Queensland, Australia*

*^9^ School of Applied Psychology, Griffith University, Queensland, Australia*

*^10^ Joint Center for Neuroscience and Neural Engineering, and Department of Biology, Southern University of Science and Technology, Shenzhen, Guangdong Province, P. R. China*

† = co-senior author

### 1 Image Acquisition

A preoperative T1-weighted MP2RAGE and a T2-weighted FLAIR sequence were obtained at baseline, using a 3T Siemens Prisma and a 64-channel head coil. The acquisition parameters were as follows: *T1*, 1 mm^3^ voxel-resolution, TR = 4000 ms, TE = 2.91 ms, flip angle = 6°, matrix size = 256 × 256, FOV = 176 × 240 × 256; *T2*, 0.9 mm^3^ voxel-resolution, TR = 5000 ms, TE = 387 ms, matrix size = 256 × 256, FOV = 192 × 256 × 256. Postoperative CT images for all participants were acquired on a Siemens Intevo, with a resolution of 0.5mm^3^.

### 2 Focal Stimulation Effects

A simulated volume of activated tissue (VAT) in each hemisphere for each participant was estimated and the dispersion of electrical charge within local structures was determined, using a recent high-resolution subcortical atlas (Pauli *et al.*, 2018). The intersection of this VAT for each structure was correlated with the percentage reduction in OCD symptoms (Supplementary Figure 1), including the terminal fibres of the stria terminalis (A), the hypothalamus (B), the nucleus accumbens (C) and ventral pallidum (D).

### 3 Cortical Connectivity

Cortical regions from the Desikan-Killiany-Tourville labelling protocol were selected based on findings from an ‘optimal’ connectivity map, derived from correlating each brain voxel (weighted by structural connectivity) to YBOCS reduction. The structural connectivity of the stimulation field with these regions was calculated and the correlation between connectivity and percentage OCD symptom reduction was visualised (Supplementary Figure 2), including the right orbitofrontal (A), right parahippocampal (B), right pars triangularis (C), right pericalcarine (D), right supramarginal (E) and right paracentral (F) regions.

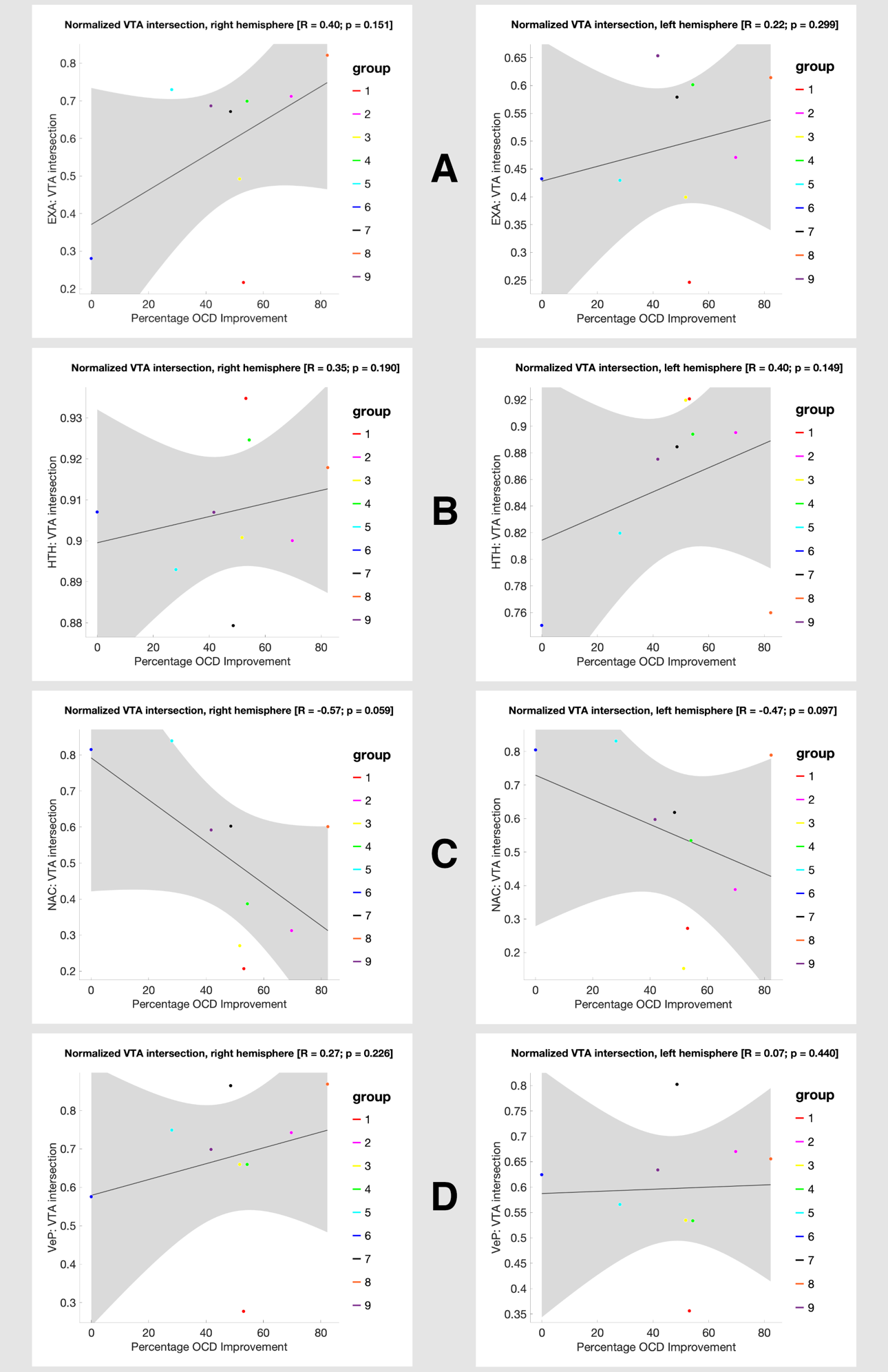

**Supplementary Figure 1 | Correlation of YBOCS Reduction with Focal Stimulation in Local Structures**

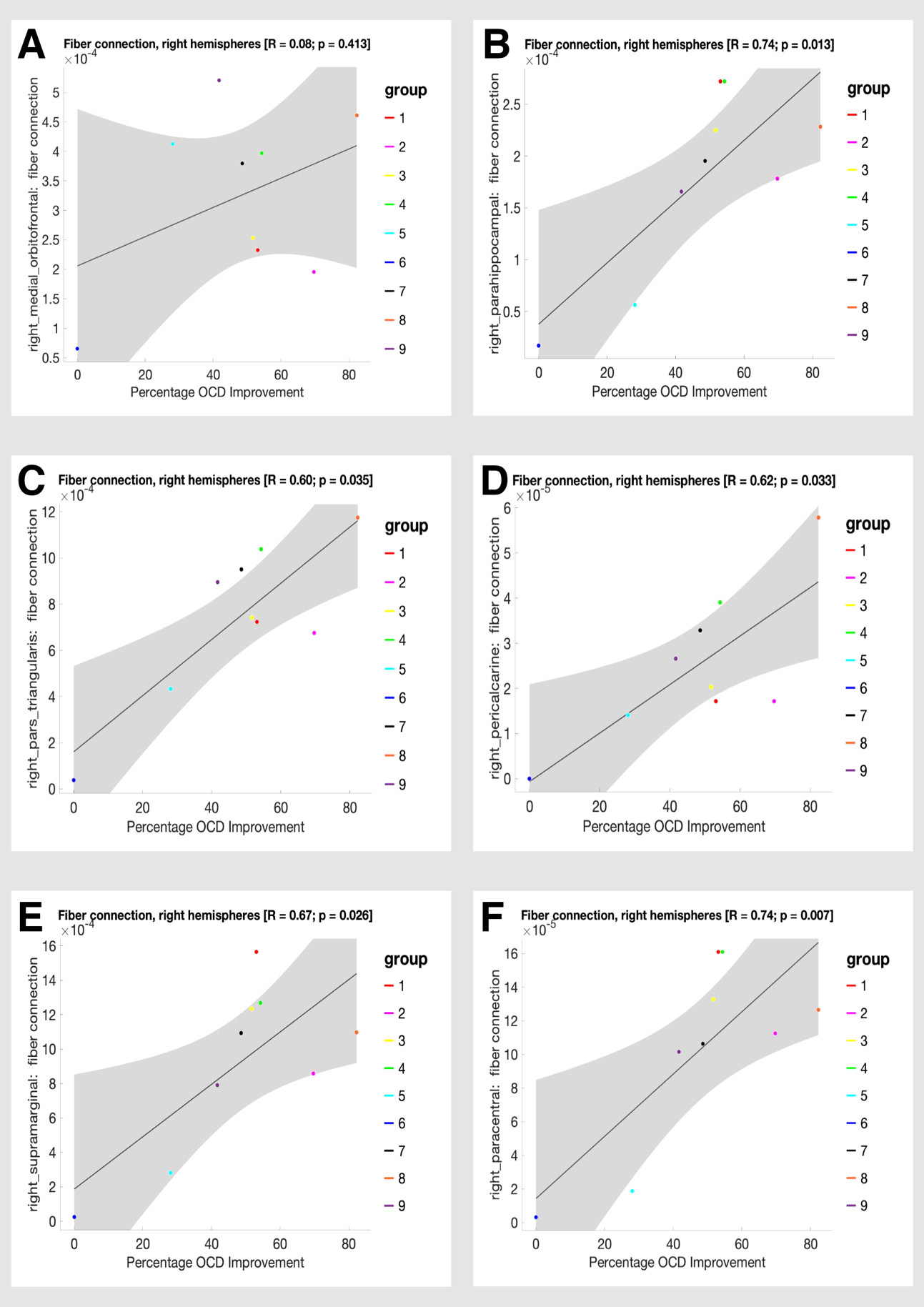

**Supplementary Figure 2 | Correlation of YBOCS Reduction with Cortical Connectivity**

**Supplementary Table 1 | Local Maxima of Cortical Regions Connected to the Stimulation Field Showing a Positive Correlation with YBOCS Reduction**

Coordinates are reported in Montreal Neurological Institute (MNI) space. R values refer to the peak voxel intensity.

| Cluster | Region | Voxel | Hemisphere | X | Y | Z | R |
| --- | --- | --- | --- | --- | --- | --- | --- |
| 1 | **Superior Orbital Gyrus (BA10)** | **161984** | **R** | **27** | **59** | **-7** | **1.10** |
|  | **Inferior Frontal Gyrus (BA 47)** |  | **R** | **40** | **43** | **-8** | **1.10** |
|  | **Medial Orbitofrontal Cortex (BA 11)** |  | **R** | **12** | **41** | **-11** | **1.10** |
|  | **Middle Frontal Gyrus (BA8)** |  | **R** | **34** | **16** | **38** | **1.10** |
|  | **Middle Temporal Gyrus (BA38)** |  | **R** | **60** | **5** | **-27** | **1.09** |
|  | **Fusiform Gyrus (BA37)** |  | **R** | **27** | **-47** | **-17** | **1.09** |
|  | **Calcarine Gyrus (BA31)** |  | **R** | **5** | **-63** | **19** | **1.08** |
| 2 | **Precentral Gyrus (BA6)** | **50** | **R** | **38** | **-19** | **67** | **0.80** |
| 3 | **Supramarginal Gyrus (BA40)** | **40** | **R** | **56** | **-15** | **24** | **0.40** |

### 4 References

Pauli, W.M., Nili, A.N., Tyszka, J.M., 2018. A high-resolution probabilistic in vivo atlas of human subcortical brain nuclei. Sci Data. 5**,** 180063.
